# Timing and Context Features in Machine Learning Classification of Inter-Patient ECG Heartbeats on the MIT-BIH Benchmark

**DOI:** 10.64898/2026.10.04.26364683

**Authors:** Edmayelle Villavicencio Alforja, Janus Capellan Aban

## Abstract

Explicit timing and context features may complement short electrocardiogram waveforms, but aggregate gains on imbalanced benchmarks can conceal errors in individual classes. We therefore compared Logistic Regression, Random Forest, XGBoost, and a one-dimensional convolutional neural network (CNN) in four cumulative input stages. Stage S1 used a 255-sample beat waveform alone, and stages S2 to S4 added 6, 13, and 21 engineered features. Using the inter-patient division of the MIT-BIH Arrhythmia Database, with 50,992 development and 49,686 evaluation beats in four classes, we estimated 95% intervals with a paired bootstrap over the 22 evaluation recordings. Adding six RR interval features increased macro F1 by 0.025 to 0.093, with intervals excluding zero for every model family. Further context helped Random Forest and XGBoost, whereas later CNN gains fell within resampling uncertainty. At the final stage, XGBoost and CNN reached macro F1 of 0.580 and 0.574, and the interval for their difference included zero. Despite opposite operating points, their precision for fusion beats was similar at matched recall, suggesting that neither separated fusion beats better. The most accurate pipeline, Random Forest at S2 with 94.45% accuracy, detected only 1.4% of supraventricular beats, and development and evaluation favored the same stage for only one family. The results for the minority classes were based on single recordings. Record 213 supplied 203 of the 207 correct CNN fusion detections, and elsewhere both final models detected at most 4 of 26 fusion beats. Record 232 contains 75% of the evaluation supraventricular beats and, nearly all were missed. Excluding this record nearly doubled XGBoost supraventricular F1, from 0.199 to 0.383. These findings are limited to retrospective analysis with annotated beat positions and a single training seed.

## 1 Introduction

Heartbeat classification assigns individual electrocardiogram beats to diagnostic categories on the basis of waveform morphology and the timing between successive beats. The MIT-BIH Arrhythmia Database is the most widely used benchmark for this task [1, 2], and its distribution through PhysioNet provides open access to the reference-annotated recordings [3]. The ANSI/AAMI EC57 standard sets out recommended practice for reporting beat-by-beat performance [4], and reviews of the field document a wide range of features, classifiers, and evaluation protocols [5].

The inter-patient protocol introduced by de Chazal et al. divides the 44 non-paced recordings into a development set (DS1) and an evaluation set (DS2) that share no records [6]. Their linear discriminant classifier, built on engineered morphology and RR-interval features, established a reference for detecting ectopic beats in unseen patients. Subsequent work selected features for generalization across patients [7], combined morphological and dynamic features [8], normalized RR intervals by subject-level statistics [9], fused temporal and morphological descriptors in ensembles of support vector machines [10], and applied random forests to small selected feature sets [11]. A recurring observation is that supraventricular and fusion beats remain difficult to detect in unseen patients [5, 18].

Convolutional neural networks (CNNs) have been applied to patient-specific classification [12] and to learning transferable heartbeat representations [13]. Hybrid methods complement these learned representations with explicit timing information: RR and frequency features have been combined with a one-dimensional CNN [14], RR intervals have been encoded as auxiliary waveforms in a multistage convolutional method [15], and RR timing has been injected into self-operational neural networks [16]. That temporal and learned features can be combined is therefore well established. Less clear is how each additional input group changes class-specific outcomes for different learners under a single, fixed evaluation protocol.

Two properties of the benchmark complicate this assessment. First, the class distribution is strongly imbalanced: aggregate scores dominated by normal beats can obscure errors in the infrequent classes, so precision and recall must be read together [17]. Second, the minority classes in MIT-BIH are concentrated in a few recordings, which limits what any single division into training and test data can reveal about generalization [18]. In addition, model selection must be kept separate from final assessment, because a score optimized during selection is an optimistically biased estimate of performance [19, 20].

In this study, we compare logistic regression, random forest, XGBoost, and a CNN across four cumulative input stages on identical beats and identical record-grouped development folds. Specifically, we quantify how aggregate performance changes as timing and context are added, characterize how class-specific performance and the trade-off between precision and recall vary across model families, and assess how sensitive these conclusions are to the choice of metric, to the split between development and evaluation data, and to the record composition of the minority classes. Since each stage was tuned independently, the stage contrasts compare trained pipelines rather than isolated feature effects. This work provides a controlled, fully reported comparison with record-level uncertainty estimates, threshold-independent precision–recall analysis, and exact per-record error accounting, which together identify what the benchmark can and cannot support.

## 2 Methods

### 2.1 Dataset and Classification Targets

The MIT-BIH Arrhythmia Database contains 48 two-channel ambulatory recordings of approximately 30 minutes each, sampled at 360 Hz and obtained from 47 subjects [1, 2]. Following de Chazal et al. [6], we used 44 recordings divided into subsets DS1 and DS2 with 22 recordings each, listed in Appendix A. Records 102, 104, 107, and 217 were excluded because they contain paced beats, consistent with the practice recommended by AAMI [4, 6].

Beats were grouped into four classes: N for non-ectopic, S for supraventricular ectopic, V for ventricular ectopic, and F for fusion of ventricular and normal beats. Annotation symbols N, L, R, e, and j were mapped to N; A, a, J, and S to S; V and E to V; and F to F. The N class therefore includes bundle branch block and escape beats and should not be read as exclusively healthy sinus beats. The unclassifiable class Q of the five-class AAMI scheme was not modeled because once the paced records are removed, it contains very few beats in either partition. This four-class grouping follows AAMI style categories without claiming compliance with every provision of the standard. Annotations outside the mapping were excluded as classification targets but retained as beat events for RR timing, so that their omission did not artificially lengthen intervals between retained beats. The retained heartbeat class distribution is given in Table 1.

**Table 1:** Retained heartbeat distribution by class and partition.

| Class | DS1 count | DS1 % | DS2 count | DS2 % |
| --- | --- | --- | --- | --- |
| N | 45,846 | 89.91 | 44,241 | 89.04 |
| S | 944 | 1.85 | 1,837 | 3.70 |
| V | 3,788 | 7.43 | 3,220 | 6.48 |
| F | 414 | 0.81 | 388 | 0.78 |
| Total | 50,992 | 100.00 | 49,686 | 100.00 |

DS1 and DS2 share no common records. However, records 201 and 202 originate from the same subject [21]. Because record 201 is in DS1 and record 202 is in DS2, the division is record disjoint rather than strictly subject disjoint.

The minority classes are concentrated in individual recordings, as summarized in Table 2. In the retained DS2 beats, record 213 contains 362 of the 388 fusion beats, or 93.3%, and record 232 contains 1,382 of the 1,837 supraventricular beats, or 75.2%. In record 232, supraventricular beats outnumber all other retained beats by more than three to one. In DS1, the database directory [23] lists 373 fusion beats in record 208, approximately 90% of the 414 retained DS1 fusion beats. The directory notes that the fusion beats in record 213 range in morphology from almost normal to almost identical to its premature ventricular contractions [22]. Consequently, F-class performance may be strongly influenced by differences in fusion-beat morphology between subjects, while DS2 S-class performance is heavily influenced by a single recording.

**Table 2:** Concentration of minority class beats in single recordings. DS2 counts are retained beats. The DS1 count is from the MIT-BIH database directory [23].

| Class | Partition | Class total | Dominant record | Beats in record | Share, % |
| --- | --- | --- | --- | --- | --- |
| F | DS1 | 414 | 208 | 373 | 90 |
| F | DS2 | 388 | 213 | 362 | 93.3 |
| S | DS2 | 1,837 | 232 | 1,382 | 75.2 |

### 2.2 Signal Preprocessing and Beat Construction

Modified limb lead II, abbreviated MLII, was selected when available, followed by lead II, and otherwise the first channel. All 44 retained recordings include an MLII channel [22], so the fallback rules were not invoked. Each signal was filtered with a fourth-order Butterworth band-pass filter from 0.5 to 40 Hz, applied forward and backward in second-order sections. Beat windows contained 128 samples before the annotated fiducial point, the fiducial sample, and 126 samples after it, for a total of 255 samples. Windows extending beyond the recording boundaries were excluded. Each retained waveform was standardized to zero mean and unit variance. When its variance fell below 10*^−^*^8^, only mean centering was applied.

This procedure yielded 100,678 retained beats. Unique beat and record identifiers maintained row alignment between waveform and feature arrays. Morphology features were computed from the standardized waveforms and therefore describe relative waveform shape rather than absolute ECG voltage.

### 2.3 Cumulative Input Stages

Model inputs were organized into four cumulative stages, described in Table 3. Each stage retained all inputs of the preceding stage and added one group of features.

**Table 3:**
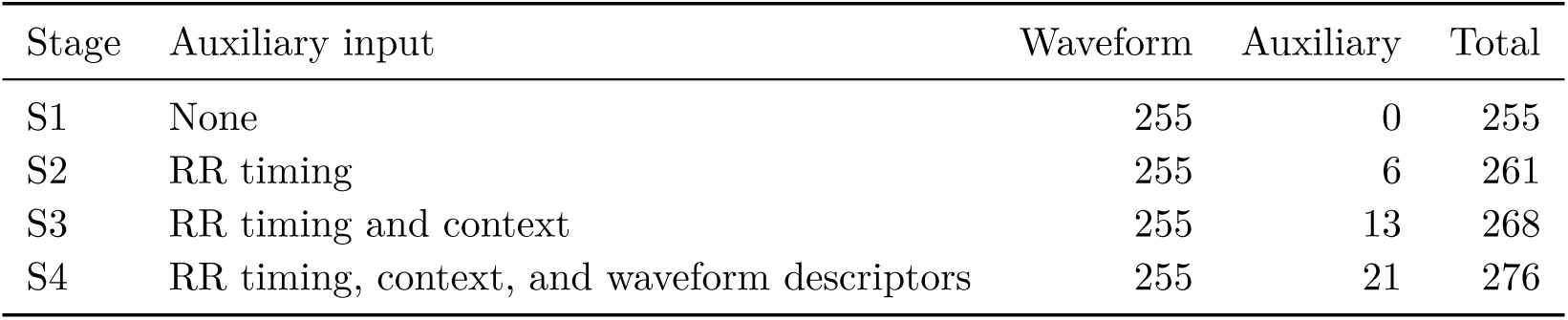
Cumulative input stages. Every stage contains the 255-sample current beat waveform.

Stage S1 contained only the current beat waveform. Stage S2 added six RR features: the preceding interval, the following interval, the median of recent historical intervals, the preceding and following intervals normalized by that median, and the ratio of the preceding to the following interval. Historical statistics used up to ten intervals completed before the current beat, excluding the immediately preceding interval, and required at least three such intervals.

Stage S3 added seven features: three rhythm history descriptors and four waveform-context descriptors. The rhythm history descriptors were the mean, standard deviation, and coefficient of variation of the historical RR intervals. The waveform-context descriptors were the Pearson correlation and the root-mean-square distance between the current waveform and each of two references, the preceding waveform and a historical waveform template. The template was the pointwise median of valid waveforms among the five preceding beat positions and required at least three valid waveforms. These seven features were added together, so their individual contributions were not isolated.

Stage S4 added eight descriptors of the current standardized waveform: minimum amplitude, maximum amplitude, peak-to-peak amplitude, mean absolute amplitude, bias corrected skewness, bias corrected excess kurtosis, mean absolute first difference, and maximum absolute first difference. Full definitions are provided in Appendix E.

### 2.4 Development and Evaluation Procedure

Five record-grouped DS1 folds were generated once and reused for all models and stages. Candidate assignments from StratifiedGroupKFold with shuffling were generated for seeds 0 to 999, and assignments lacking any class in a validation fold were rejected. The selected assignment, corresponding to seed 186, minimized the sum of the coefficient of variation of fold sizes and the mean, across classes, of the standard deviation of fold class proportions. No classifier score was used to choose the folds, and no recording was split between training and validation. Median imputation and feature scaling were fitted on each training fold and applied to its validation fold. For final evaluation, they were refitted on all of DS1 before being applied to DS2. DS2 labels were not used in any fitting or selection step.

For the classical models, one configuration per stage was selected by mean validation macro F1, with mean balanced accuracy as a secondary criterion. For the CNN, a checkpoint was selected within each fold by validation macro F1, with balanced accuracy as a tie-breaker, and the median of the five selected epochs set the duration of final DS1 training. Since each stage was optimized independently, hyperparameters, CNN training duration, and CNN parameter count could vary across stages. Comparisons between stages therefore reflect differences between complete trained pipelines rather than the isolated effects of individual feature additions. Cross-validation scores were used for model selection and should be interpreted as development scores rather than independent estimates of generalization performance [19, 20].

### 2.5 Classical Classifiers

Logistic Regression used median imputation, standard scaling, balanced class weights, the L-BFGS solver, and at most 3,000 iterations. The inverse regularization strength *C* was selected from the set *{*0.01, 0.1, 1, 10*}*. Random Forest [24] used 300 trees, balanced subsample class weighting, and median imputation without scaling. Maximum depth was selected from *{*10, 20, unrestricted*}*, and minimum leaf size was selected from *{*1, 2*}*.

XGBoost [25] used 300 estimators, maximum depth five, learning rate 0.05, row and column subsampling of 0.8, minimum child weight two, *γ* = 0.1, histogram tree construction, and a four-class soft-probability objective. Training sample weights were *w_c_* = [*n/*(4*n_c_*)]*^α^*, where *n* is the training-set size and *n_c_* the count of class *c*, with the exponent *α* selected from the set *{*0.85, 1.00, 1.15, 1.30*}*. This exponent controls class weighting and is distinct from XGBoost’s L1 regularization parameter. Class predictions were taken as the most probable class.

Search budgets differed across families: four configurations for Logistic Regression, six for Random Forest, four for XGBoost, and only the training epoch for the CNN. Several selected values lay at the lower boundary of their grids: *C* = 0.01 at S1 and S2, maximum depth 10 at S1, S3, and S4, and *α* = 0.85 at S1, S3, and S4. Better-performing settings may therefore lie outside the searched ranges. All selected settings are listed in Appendix Table A.1.

### 2.6 Convolutional Neural Network Classifier

The CNN used a common waveform backbone at every stage: three convolutional blocks with 32, 64, and 128 channels and kernel sizes 7, 5, and 3, each followed by batch normalization and ReLU activation, with max pooling by a factor of 2 after the first two blocks. Adaptive average pooling produced a 128-dimensional waveform representation.

For stages with auxiliary inputs, a feature branch mapped the auxiliary vector through layers of 64 and 32 units with ReLU activation and dropout of 0.20 after the first layer. The classifier concatenated the waveform representation, the learned auxiliary representation, and the original preprocessed auxiliary vector, and passed them through a 128-unit hidden layer with ReLU and dropout of 0.30 to four output logits. The waveform-only stage omitted the auxiliary branch. Parameter counts were 52,740, 60,132, 61,476, and 63,012 for S1 to S4, so the stage comparison also changes model capacity.

The network was implemented in PyTorch [26] and trained with Adam [27] at a learning rate of 0.001 and a batch size of 256, using class-weighted cross-entropy with balanced weights computed within each training fold. Each fold was trained for 20 epochs, retaining the best validation checkpoint. Final training used 14, 10, 7, and 5 epochs for S1 to S4, respectively. The decline in selected epochs as features were added is consistent with earlier overfitting in richer stages. Because the final model was trained on all of DS1, about 25% more data than each fold model, the median fold epoch is an approximation to the optimal final duration. Auxiliary features were median-imputed and standardized using training-only estimates. Classical estimators used random seed 42, CNN fold training used seed 42 + *k* for fold *k ∈ {*1*, … ,* 5*}*, and final training used seed 42, with Python, NumPy, and PyTorch generators seeded. Class imbalance was addressed by weighting only, without resampling.

### 2.7 Outcomes and Satistical Scope

The primary outcome was DS2 macro F1, the unweighted mean of the four class F1 scores:

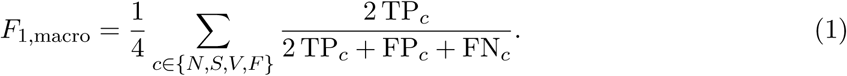

Secondary outcomes were accuracy, balanced accuracy, defined as the mean of the four class recalls, macro precision, support-weighted F1, and the multiclass Matthews correlation coefficient, MCC [28]. For each class, precision was TP*/*(TP + FP) and recall was TP*/*(TP + FN); these correspond to the positive predictivity, +P, and the sensitivity, Se, reported in much of the ECG literature. Undefined precision or F1 values were set to zero. Class decisions used the default most-probable-class rule. Signal processing and distribution-shape features used SciPy [29], while classical models and metrics used scikit-learn [30]. Stage differences were computed from unrounded scores.

To quantify uncertainty and attribute errors, the 16 saved final models were reloaded without retraining, and their DS2 predictions and class probabilities were exported together with each beat’s record identifier. The reloaded models reproduced every reported per-class precision, recall, and F1 to within 5 *×* 10*^−^*^7^. Uncertainty was estimated with a paired bootstrap over recordings: the 22 DS2 recordings were resampled with replacement 5,000 times, all beats of each sampled recording were included, and every metric was recomputed for all 16 conditions on the same resamples. Percentile 95% intervals are reported for macro F1 and for paired differences between conditions. Because whole recordings are resampled, the intervals reflect between-patient variability in the evaluation set; they do not include training variability, which would require repeated training with different seeds. Threshold-independent performance was summarized for each class with one-versus-rest precision–recall curves built from the class probabilities, their average precision, and the precision attainable at a matched recall. Per-record detections and false positives were tabulated for the S and F classes, and a leave-one-recording-out analysis recomputed DS2 metrics without record 213 or record 232.

## 3 Results

### 3.1 Aggregate Performance Across Input Stages

Figure 1 and Table 4 summarize DS2 performance for all 16 model–stage conditions. Relative to waveform-only pipelines, adding six RR features increased macro F1 by 0.0912 for Logistic Regression, 0.0246 for Random Forest, 0.0804 for XGBoost, and 0.0935 for the CNN. Adding rhythm-history and waveform-context features at S3 increased it by a further 0.0439, 0.0410, 0.0540, and 0.0345, respectively. Both transitions were positive for every family.

**Figure 1:**
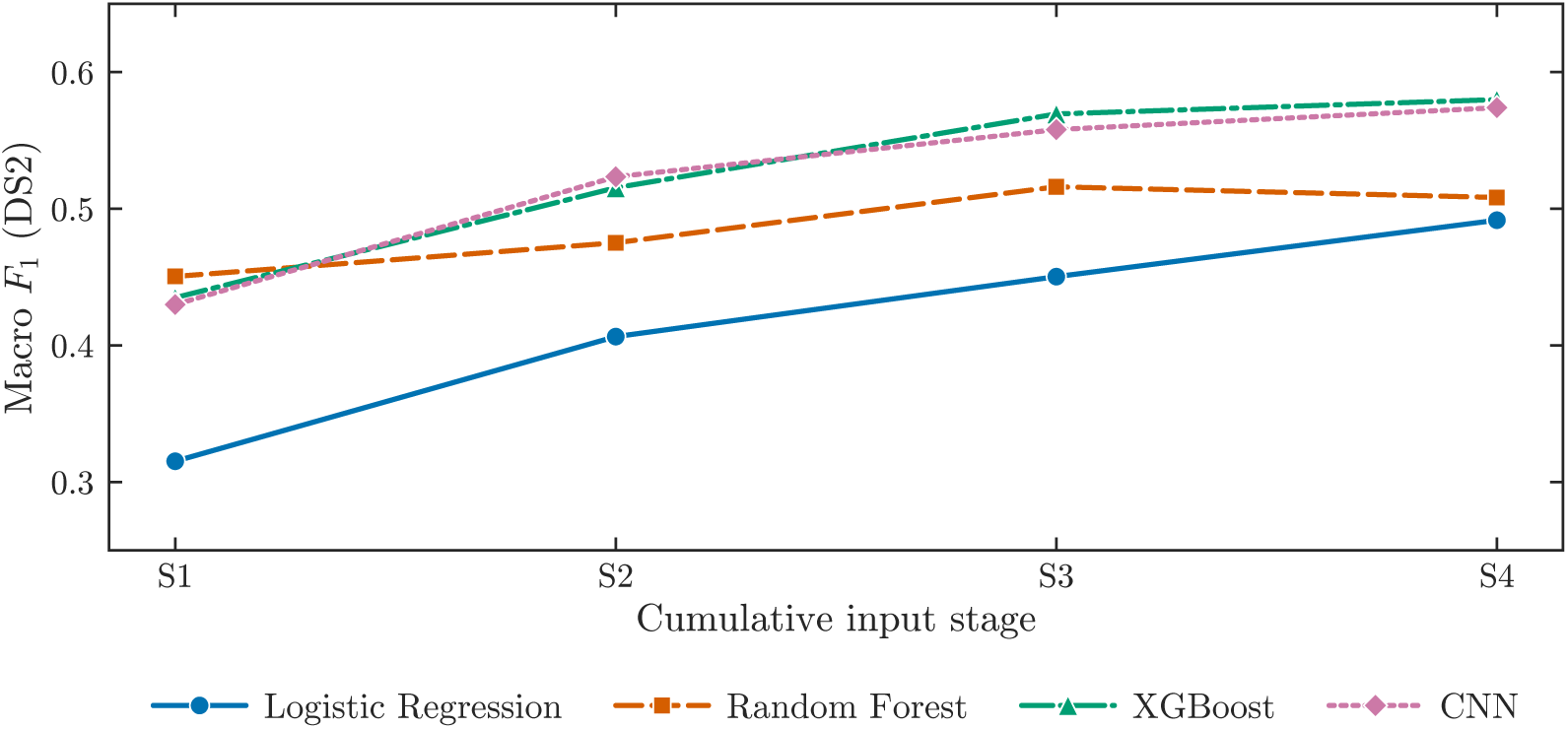
DS2 macro F1 across four cumulative input stages: S1 with the waveform only, S2 with 6 RR features, S3 with 13 timing and context features, and S4 with 21 features. Lines connect separately tuned pipelines.

**Table 4:**
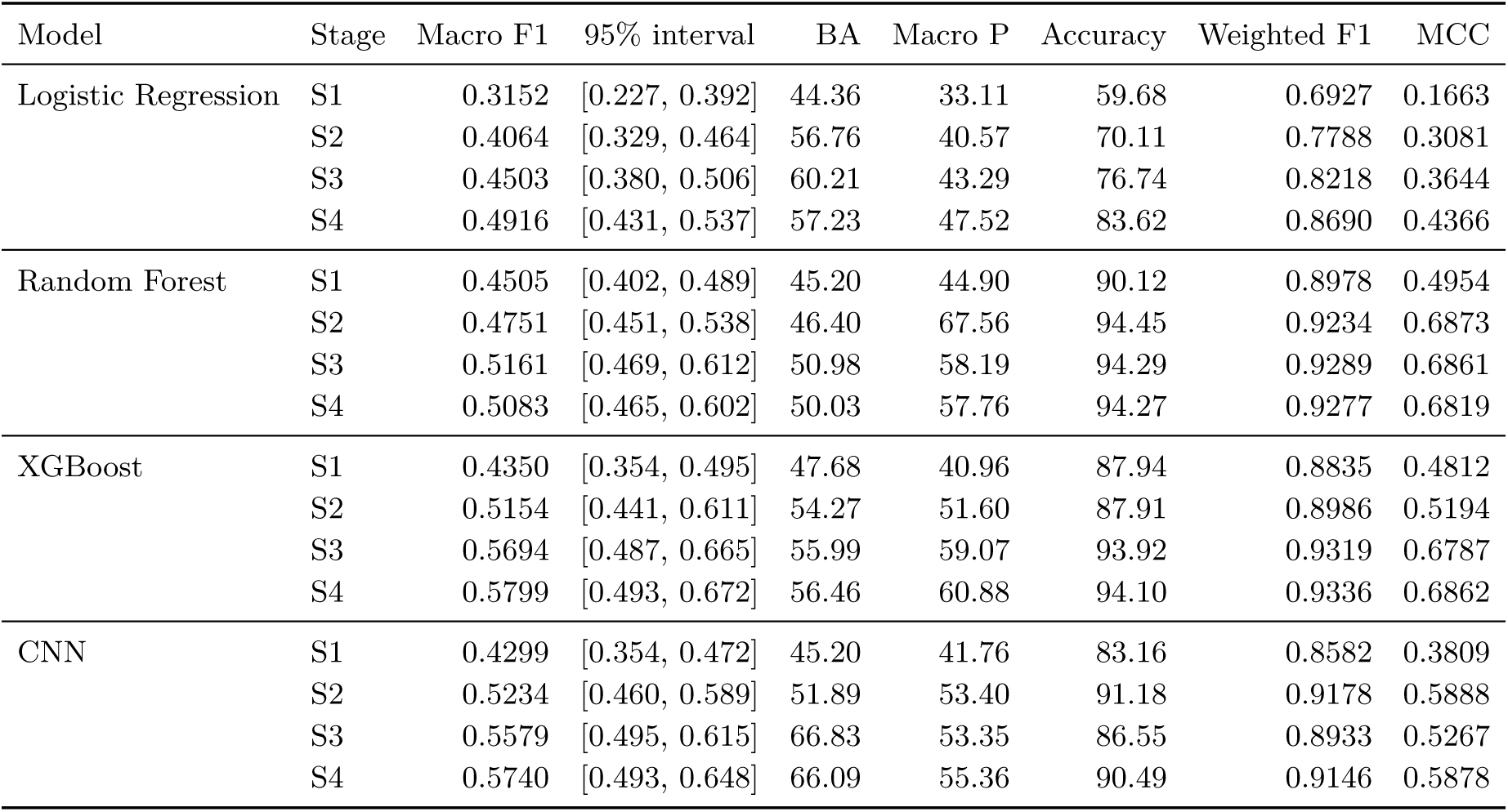
DS2 aggregate performance across all model and stage combinations. The 95% interval for macro F1 is from the record-level bootstrap. BA denotes balanced accuracy, while macro P denotes macro precision. Both metrics, together with accuracy, are reported as percentages. Weighted F1 is weighted according to class support, and MCC denotes the multiclass Matthews correlation coefficient. For reference, assigning every DS2 beat to class N would yield 89.04% accuracy, 25.00% balanced accuracy, and a macro F1 of 0.2355.

| Model | Stage | Macro F1 | 95% interval | BA | Macro P | Accuracy | Weighted F1 | MCC |
| --- | --- | --- | --- | --- | --- | --- | --- | --- |
| Logistic Regression | S1 | 0.3152 | [0.227, 0.392] | 44.36 | 33.11 | 59.68 | 0.6927 | 0.1663 |
|  | S2 | 0.4064 | [0.329, 0.464] | 56.76 | 40.57 | 70.11 | 0.7788 | 0.3081 |
|  | S3 | 0.4503 | [0.380, 0.506] | 60.21 | 43.29 | 76.74 | 0.8218 | 0.3644 |
|  | S4 | 0.4916 | [0.431, 0.537] | 57.23 | 47.52 | 83.62 | 0.8690 | 0.4366 |
| Random Forest | S1 | 0.4505 | [0.402, 0.489] | 45.20 | 44.90 | 90.12 | 0.8978 | 0.4954 |
|  | S2 | 0.4751 | [0.451, 0.538] | 46.40 | 67.56 | 94.45 | 0.9234 | 0.6873 |
|  | S3 | 0.5161 | [0.469, 0.612] | 50.98 | 58.19 | 94.29 | 0.9289 | 0.6861 |
|  | S4 | 0.5083 | [0.465, 0.602] | 50.03 | 57.76 | 94.27 | 0.9277 | 0.6819 |
| XGBoost | S1 | 0.4350 | [0.354, 0.495] | 47.68 | 40.96 | 87.94 | 0.8835 | 0.4812 |
|  | S2 | 0.5154 | [0.441, 0.611] | 54.27 | 51.60 | 87.91 | 0.8986 | 0.5194 |
|  | S3 | 0.5694 | [0.487, 0.665] | 55.99 | 59.07 | 93.92 | 0.9319 | 0.6787 |
|  | S4 | 0.5799 | [0.493, 0.672] | 56.46 | 60.88 | 94.10 | 0.9336 | 0.6862 |
| CNN | S1 | 0.4299 | [0.354, 0.472] | 45.20 | 41.76 | 83.16 | 0.8582 | 0.3809 |
|  | S2 | 0.5234 | [0.460, 0.589] | 51.89 | 53.40 | 91.18 | 0.9178 | 0.5888 |
|  | S3 | 0.5579 | [0.495, 0.615] | 66.83 | 53.35 | 86.55 | 0.8933 | 0.5267 |
|  | S4 | 0.5740 | [0.493, 0.648] | 66.09 | 55.36 | 90.49 | 0.9146 | 0.5878 |

The eight current-waveform descriptors at S4 changed macro F1 by +0.0413, *−*0.0078, +0.0105, and +0.0161 for Logistic Regression, Random Forest, XGBoost, and the CNN, respectively. Relative to S1, final stage macro F1 increased by 0.1764, 0.0578, 0.1449, and 0.1440, respectively. Random Forest led at S1, the CNN at S2, and XGBoost at S3 and S4.

Table 5 gives paired bootstrap intervals for these changes. The gain from adding RR features, from S1 to S2, was the only change whose interval excluded zero for all four families. Adding context, from S2 to S3, produced intervals excluding zero for Random Forest and XGBoost, a borderline interval for Logistic Regression, and an interval spanning zero for the CNN. Adding the waveform descriptors, from S3 to S4, produced an interval excluding zero only for Logistic Regression. At S4, XGBoost exceeded Random Forest with an interval excluding zero, whereas the XGBoost–CNN difference of 0.006 had an interval from *−*0.048 to +0.071; the evaluation set cannot rank these two pipelines.

**Table 5:** Paired record-level bootstrap contrasts in DS2 macro F1, from 5,000 resamples of the 22 DS2 recordings. *P* (*>* 0) is the proportion of resamples in which the difference was positive. The ^∗^95% interval excludes zero.

| Model or comparison | Change | Difference | 95% interval | $P(> 0)$ |
| --- | --- | --- | --- | --- |
| Logistic Regression | S1 → S2 | +0.091* | [+0.042, +0.144] | 1.00 |
| | S2 → S3 | +0.044 | [−0.003, +0.092] | 0.97 |
| | S3 → S4 | +0.041* | [+0.011, +0.076] | 1.00 |
| Random Forest | S1 → S2 | +0.025* | [+0.002, +0.089] | 0.98 |
| | S2 → S3 | +0.041* | [+0.003, +0.108] | 0.98 |
| | S3 → S4 | −0.008 | [−0.016, +0.003] | 0.09 |
| XGBoost | S1 → S2 | +0.080* | [+0.041, +0.155] | 1.00 |
| | S2 → S3 | +0.054* | [+0.014, +0.077] | 1.00 |
| | S3 → S4 | +0.011 | [−0.001, +0.027] | 0.96 |
| CNN | S1 → S2 | +0.093* | [+0.037, +0.183] | 1.00 |
| | S2 → S3 | +0.035 | [−0.065, +0.119] | 0.67 |
| | S3 → S4 | +0.016 | [−0.029, +0.064] | 0.73 |
| XGBoost − CNN | S4 | +0.006 | [−0.048, +0.071] | 0.62 |
| XGBoost − Random Forest | S4 | +0.072* | [+0.011, +0.096] | 1.00 |
| CNN − Random Forest | S4 | +0.066 | [−0.032, +0.124] | 0.89 |

The choice of metric changed the ranking. Random Forest at S2 had the highest accuracy, 94.45%, and the highest macro precision, 67.56%, but its macro F1 was 0.4751 and its balanced accuracy 46.40%. The CNN at S3 had the highest balanced accuracy, 66.83%, with an accuracy of only 86.55%. Final stage XGBoost had the highest macro F1, 0.5799, and the highest weighted F1, 0.9336, whereas the highest MCC, 0.6873, again belonged to Random Forest at S2. Four metrics thus identified three different best pipelines.

At the final stage, XGBoost and CNN macro F1 differed by only 0.0059. XGBoost had higher macro precision at 60.88% against 55.36%, higher accuracy at 94.10% against 90.49%, higher weighted F1 at 0.9336 against 0.9146, and higher MCC at 0.6862 against 0.5878, whereas the CNN had higher balanced accuracy at 66.09% against 56.46%. These differences reflect distinct operating points rather than a consistent ordering, and the bootstrap interval for their macro F1 difference spans zero.

### 3.2 Class Specific performance and operating points

Appendix Tables C.1 to C.4 report precision, recall, F1, correct and predicted counts, and support for every class and condition. Every condition was evaluated on 44,241 class N, 1,837 class S, 3,220 class V, and 388 class F beats. Figure 2 shows class F1 across stages for each family.

**Figure 2:**
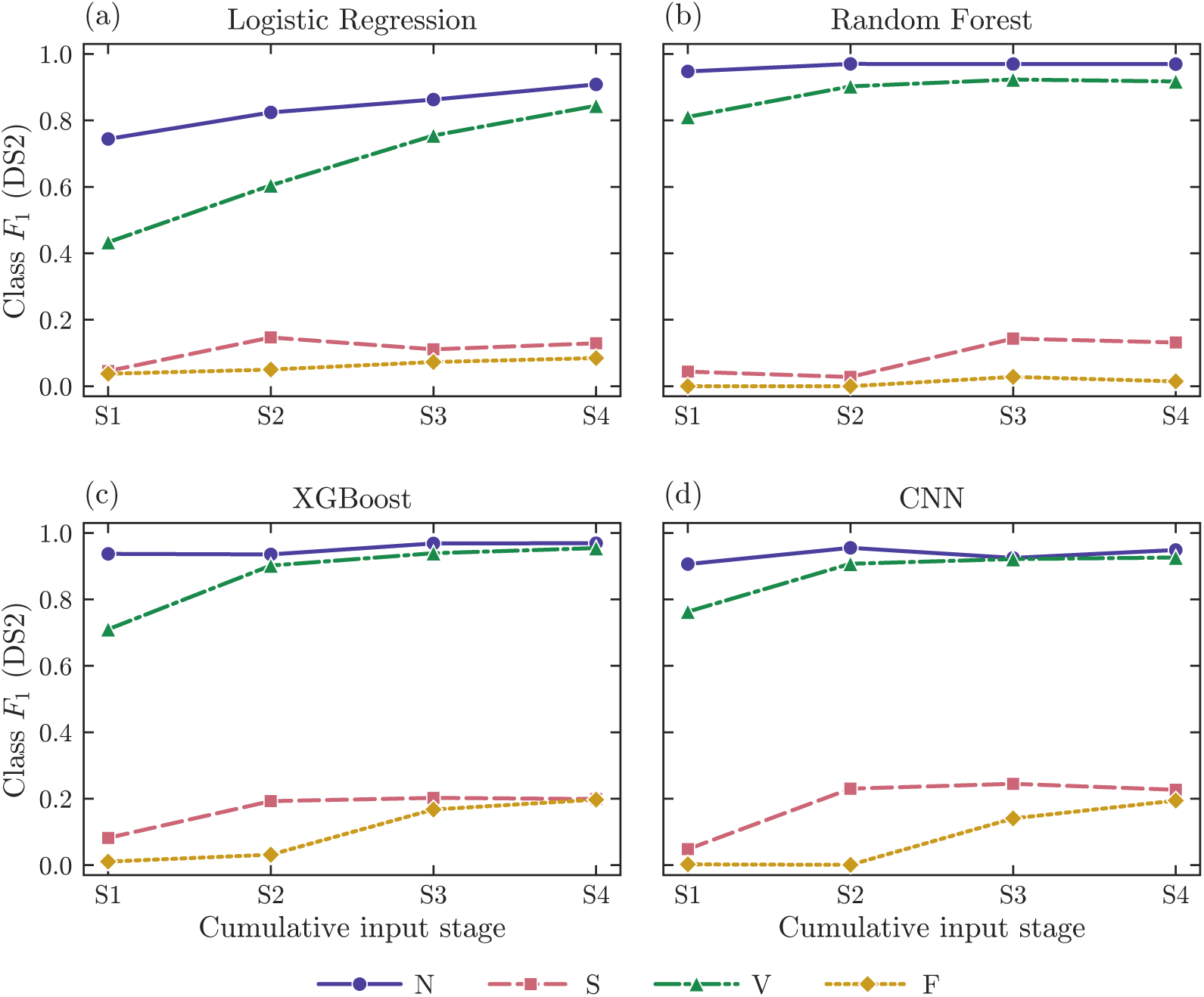
Class specific F1 on DS2 across stages S1 to S4 for each model family. Class V F1 rises with added inputs for every model family except Random Forest at S4. Class S F1 remains below 0.25 throughout, while class F F1 remains below 0.20. For Random Forest, class F F1 is near zero at every stage. Lines connect separately tuned pipelines.

For Logistic Regression, the F1 score for class N increased from 0.7442 at S1 to 0.9084 at S4, while class V F1 increased from 0.4335 to 0.8442. Class S F1 peaked at 0.1468 at S2, decreased to 0.1110 at S3, and recovered to 0.1292 at S4. Class F F1 also increased across stages, from 0.0376 at S1 to 0.0847 at S4, although precision remained below 5% throughout. From S3 to S4, class F recall decreased from 56.44% to 33.76%, while precision increased from 3.90% to 4.84%. Thus, the increase in class F F1 did not correspond to an increase in sensitivity.

Random Forest performance was concentrated in classes N and V. At S2, recall reached 99.88% for class N and 84.29% for class V. In contrast, the model predicted only 33 beats as class S, of which 26 were correct, yielding a recall of 1.42%. It predicted only one beat as class F, and that prediction was incorrect. At S1, the model predicted 185 beats as class F, but none were correct. At S3, class S F1 reached 0.1433, while class F F1 reached 0.0281, with 6 of 388 F beats correctly detected. At S4, these values decreased to 0.1316 for class S and 0.0146 for class F, with only 3 F beats correctly detected. Class V F1 also decreased slightly, from 0.9231 at S3 to 0.9174 at S4. Thus, the high overall accuracy of Random Forest coexisted with very poor performance on the two minority classes.

For XGBoost, the F1 scores for classes V and F increased at every stage, reaching 0.9543 and 0.1972, respectively, at S4. Class S F1 peaked at 0.2025 at S3 and remained similar at 0.1991 at S4. At S4, the model predicted 996 beats as class S, of which 282 were correct, corresponding to 28.31% precision and 15.35% recall. It also predicted 261 beats as class F, of which 64 were correct, yielding 24.52% precision and 16.49% recall.

For the CNN, performance gains were uneven across classes. Class V F1 increased at every stage, from 0.7630 at S1 to 0.9260 at S4. Class N F1 peaked at 0.9552 at S2, decreased at S3, and partially recovered at S4. Class S F1 increased from 0.0481 at S1 to 0.2302 at S2, peaked at 0.2449 at S3, and declined slightly to 0.2269 at S4. Class F F1 remained near zero at S1 and S2, then increased to 0.1407 at S3 and 0.1945 at S4. From S2 to S3, class S recall increased from 19.00% to 37.67%, while precision decreased from 29.21% to 18.14%. At S4, the CNN predicted 1,741 beats as class F, of which 207 were correct, corresponding to 11.89% precision and 53.35% recall. Thus, fewer than one in eight F predictions corresponded to a true fusion beat.

XGBoost and the CNN achieved similar class F F1 scores at S4, 0.1972 and 0.1945, respectively, but through different precision and recall trade-offs. The CNN detected 207 of 388 fusion beats, compared with 64 for XGBoost, but its precision was less than half that of XGBoost. For class S, the CNN also achieved higher recall, 22.16% against 15.35%, but lower precision, 23.24% against 28.31%, resulting in F1 scores of 0.2269 and 0.1991, respectively. False-positive counts refer to beat-level assignments, while alarm rates per recording or per unit time were not evaluated.

Figure 3 places all 16 conditions in precision–recall space for each class. For class N, every condition has precision between 0.93 and 0.97, so the families differ almost entirely in N recall, from 0.62 for Logistic Regression at S1 to 0.999 for Random Forest at S2. For class V, added inputs raised precision more than recall: Logistic Regression V precision rose from 0.34 at S1 to 0.80 at S4, and XGBoost V precision from 0.59 to 0.95, while the recall of most families was already above 0.8 at S1. For class S, the families occupy different regions: Random Forest at high precision and very low recall, Logistic Regression at low precision, and XGBoost and the CNN between them, with no condition exceeding an F1 of 0.25. For class F, the CNN and Logistic Regression reach recall above 0.3 only at precision below 0.12, whereas XGBoost moves toward higher precision at low recall. The points trace distinct trade-offs rather than a single frontier along which one family dominates.

**Figure 3:**
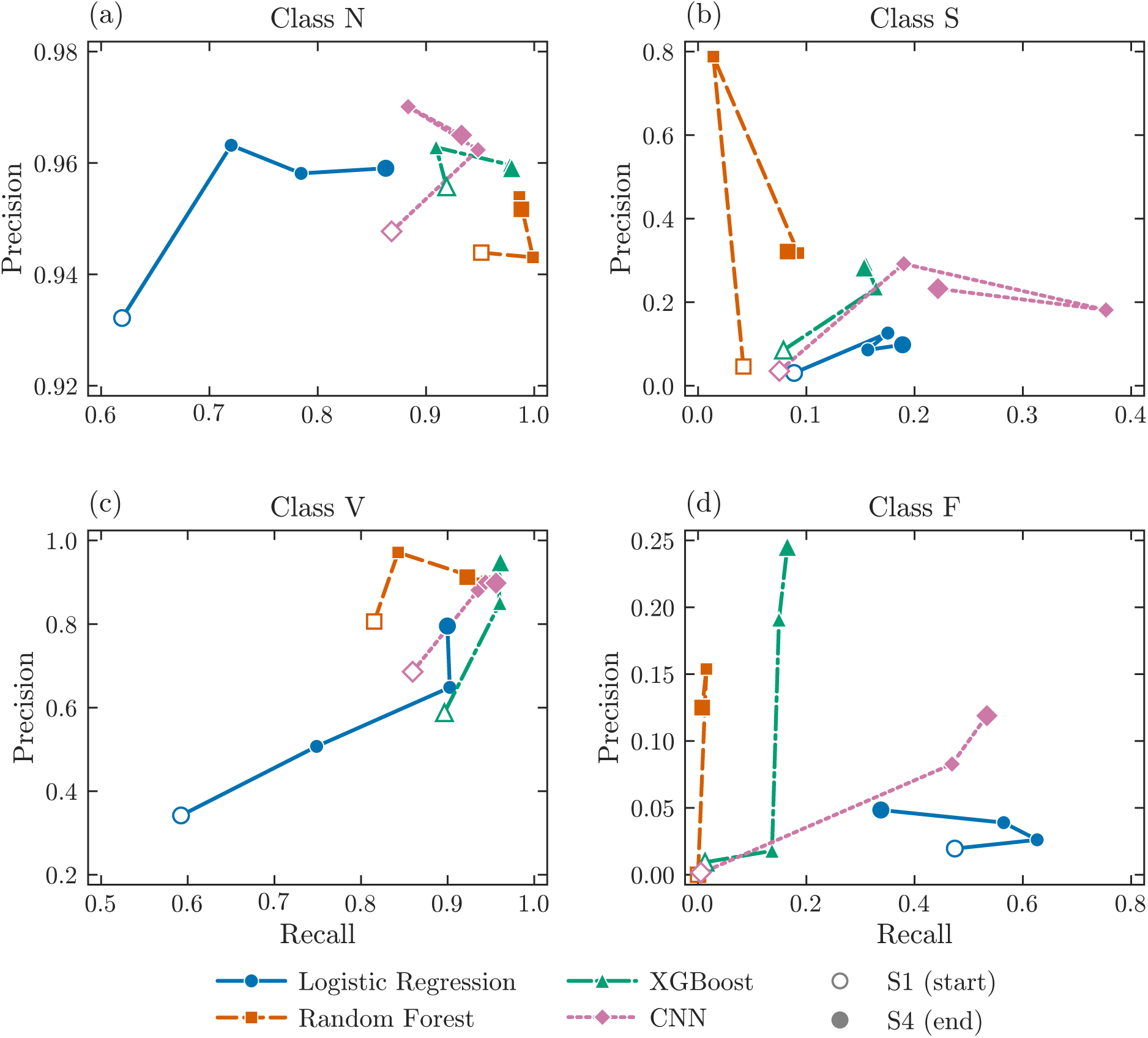
DS2 operating points for each class. Each line traces one model family from S1 to S4. Each point represents the operating point produced by the default most probable class rule.

Threshold-independent analysis shows that these trade-offs come mainly from the decision rule. Figure 4 shows the final-stage precision–recall curves, and Table 6 gives the precision each model attains at matched recall. For class F, the CNN and XGBoost had similar average precision, 15.1% and 13.9%. At XGBoost’s default recall of 16.5%, the CNN’s curve reached 28.4% precision against 23.3% for XGBoost, and at the CNN’s recall of 53.4%, both curves gave about 12% precision. For class S, the two models were again close at matched recall, with about 29% precision at 15% recall and 22 to 23% at 22% recall, although the CNN had higher average precision, 20.3% against 14.7%. The opposite operating points of the two models therefore arise largely from where their default decisions fall on similar curves, which reflects calibration and class weighting rather than better learned representations. Logistic Regression and Random Forest were well below both on the minority classes at every recall.

**Figure 4:**
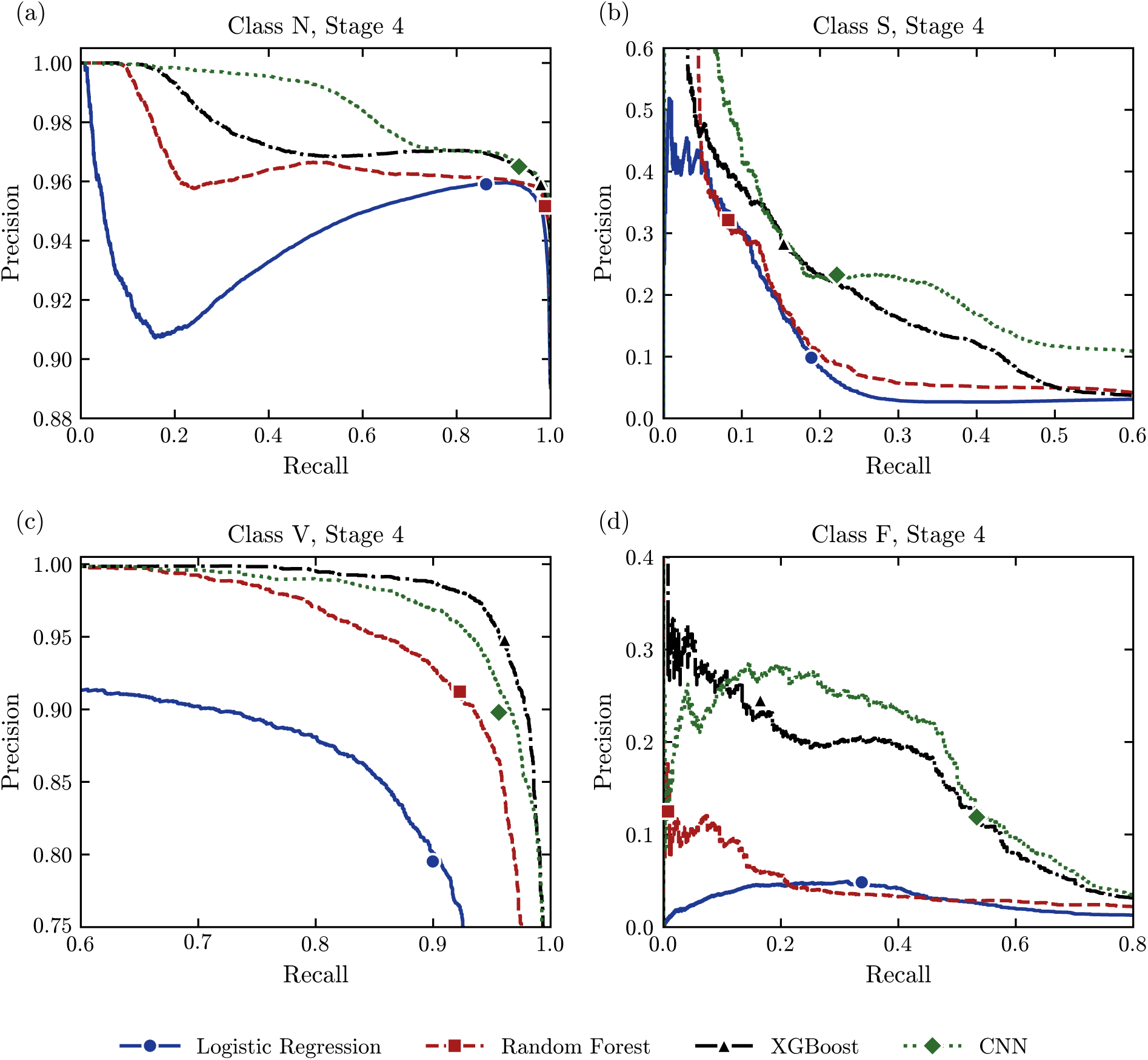
Precision and recall curves on DS2 for the final stage S4 models, computed from each model’s class probabilities. Filled markers indicate the default operating point produced by the most probable class rule.

**Table 6:** Threshold independent comparison of the final stage S4 models for the minority classes, reported in percentage. Precision at matched recall is defined as the highest precision achieved on each model’s precision and recall curve at or above the default recall of the reference model. Average precision is the area under the precision-recall curve and summarizes performance across all thresholds.

| Class | Quantity | LR | RF | XGBoost | CNN |
| --- | --- | --- | --- | --- | --- |
| S | Precision at XGBoost recall, 15.4% | 16.7 | 17.7 | 29.1 | 28.9 |
| S | Precision at CNN recall, 22.2% | 5.9 | 8.8 | 22.1 | 23.4 |
| S | Average precision | 8.5 | 11.1 | 14.7 | 20.3 |
| F | Precision at XGBoost recall, 16.5% | 5.0 | 6.0 | 23.3 | 28.4 |
| F | Precision at CNN recall, 53.4% | 2.6 | 2.9 | 11.9 | 12.9 |
| F | Average precision | 2.7 | 4.0 | 13.9 | 15.1 |

### 3.3 Record Level results

The Table 7 lists for each DS2 recording the S and F beats detected and the false positives produced by final-stage XGBoost and the CNN. Minority class performance depends almost entirely on two recordings.

**Table 7:** Record level detections and false positives for classes S and F on DS2 using the final stage XGBoost and CNN models. The Det. denotes the number of correctly detected beats, FP denotes false positives, and XGB denotes XGBoost. True denotes the number of annotated beats of the corresponding class in each recording. Records 213 and 232 are shown in bold.

| Record | S class |  |  |  |  | F class |  |  |  |  |
| --- | --- | --- | --- | --- | --- | --- | --- | --- | --- | --- |
|  | True | XGB |  | CNN |  | True | XGB |  | CNN |  |
|  |  | Det. | FP | Det. | FP |  | Det. | FP | Det. | FP |
| 100 | 33 | 33 | 1 | 30 | 1 | 0 | 0 | 0 | 0 | 2 |
| 103 | 2 | 0 | 0 | 1 | 7 | 0 | 0 | 0 | 0 | 3 |
| 105 | 0 | 0 | 16 | 0 | 31 | 0 | 0 | 10 | 0 | 238 |
| 111 | 0 | 0 | 0 | 0 | 2 | 0 | 0 | 0 | 0 | 31 |
| 113 | 6 | 4 | 0 | 6 | 48 | 0 | 0 | 0 | 0 | 0 |
| 117 | 1 | 1 | 0 | 0 | 0 | 0 | 0 | 0 | 0 | 0 |
| 121 | 1 | 0 | 0 | 1 | 22 | 0 | 0 | 0 | 0 | 19 |
| 123 | 0 | 0 | 3 | 0 | 6 | 0 | 0 | 0 | 0 | 0 |
| 200 | 30 | 20 | 21 | 26 | 47 | 2 | 0 | 1 | 1 | 315 |
| 202 | 55 | 29 | 4 | 35 | 175 | 1 | 1 | 5 | 0 | 230 |
| 210 | 22 | 2 | 22 | 9 | 160 | 10 | 0 | 8 | 2 | 175 |
| 212 | 0 | 0 | 0 | 0 | 2 | 0 | 0 | 0 | 0 | 11 |
| **213** | 28 | 0 | 0 | 8 | 0 | 362 | 63 | 21 | 203 | 57 |
| 214 | 0 | 0 | 18 | 0 | 32 | 1 | 0 | 3 | 0 | 20 |
| 219 | 7 | 4 | 384 | 4 | 425 | 1 | 0 | 0 | 0 | 18 |
| 221 | 0 | 0 | 2 | 0 | 105 | 0 | 0 | 0 | 0 | 36 |
| 222 | 209 | 172 | 239 | 180 | 256 | 0 | 0 | 3 | 0 | 127 |
| 228 | 3 | 3 | 2 | 2 | 5 | 0 | 0 | 0 | 0 | 2 |
| 231 | 1 | 0 | 2 | 0 | 18 | 0 | 0 | 0 | 0 | 0 |
| **232** | 1,382 | 5 | 0 | 91 | 0 | 0 | 0 | 0 | 0 | 1 |
| 233 | 7 | 6 | 0 | 4 | 1 | 11 | 0 | 146 | 1 | 221 |
| 234 | 50 | 3 | 0 | 10 | 1 | 0 | 0 | 0 | 0 | 28 |
| Total | 1,837 | 282 | 714 | 407 | 1,344 | 388 | 64 | 197 | 207 | 1,534 |

Concerning fusion beats Record 213 supplied 63 of XGBoost’s 64 and 203 of the CNN’s 207 correct F detections. Outside that recording, the two models detected 1 and 4 of the remaining 26 fusion beats. Their difference in F sensitivity is therefore a difference on one patient, where XGBoost detected 17.4% and the CNN 56.1% of 362 fusion beats. The false positives behaved differently: 146 of XGBoost’s 197 false F predictions came from record 233, whereas the CNN’s 1,534 were spread over 18 recordings, with no single recording contributing more than 21%. Without record 213, F-class F1 fell to 0.010 for XGBoost and 0.005 for the CNN.

In addition for supraventricular beats Record 232 contains 75% of DS2 S beats but was almost entirely missed: XGBoost detected 5 and the CNN 91 of its 1,382 S beats, and Random Forest detected 4. In the other recordings, XGBoost detected 277 and the CNN 316 of the 455 S beats, including 172 and 180 of the 209 in record 222. Most S false positives came from two recordings, 219 and 222, which together produced 623 of XGBoost’s 714 and 681 of the CNN’s 1,344. Excluding record 232 raised S-class F1 from 0.199 to 0.383 for XGBoost, from 0.227 to 0.299 for the CNN, and from 0.132 to 0.320 for Random Forest, and raised XGBoost macro F1 from 0.580 to 0.630.

### 3.4 Agreement Between Development and Evaluation

Mean DS1 development macro F1 favored S4 for Logistic Regression and Random Forest but S3 for XGBoost and the CNN, as shown in Appendix Table B.1. Table 8 reports the DS2 outcomes of these development-preferred stages. On DS2, the highest macro F1 occurred at S4 for Logistic Regression, S3 for Random Forest, and S4 for XGBoost and the CNN. Development and evaluation therefore agreed on the best stage only for Logistic Regression. The DS2 rankings are reported descriptively and do not replace the development-based selections. Moreover, a mean of fold-wise F1 scores and an F1 computed on the pooled DS2 population are different aggregations, so their difference is not a direct estimate of a generalization gap.

**Table 8:** DS2 outcomes for the stage with the highest mean DS1 development macro F1 for each model. Development scores are selection dependent and shown for context.

| Model | Selected stage | DS1 macro F1 | DS2 macro F1 | DS2 balanced accuracy |
| --- | --- | --- | --- | --- |
| Logistic Regression | S4 | 0.4022 | 0.4916 | 0.5723 |
| Random Forest | S4 | 0.4436 | 0.5083 | 0.5003 |
| XGBoost | S3 | 0.4679 | 0.5694 | 0.5599 |
| CNN | S3 | 0.5528 | 0.5579 | 0.6683 |

The fold composition helps explain this instability. Although the fold sizes were similar, validation fold 5 contained 372 of the 414 DS1 F beats, accounting for 89.9% of the total, because record 208 was assigned to that fold. This left only 42 F beats available for training in fold 5. Fold 2 contained only four F validation beats, while fold 5 contained only five S validation beats, as shown in Appendix Table B.2. F1 scores calculated from folds with so few minority-class examples are highly variable. The fold standard deviations reported in Appendix Table B.1 therefore reflect differences in class composition across folds rather than confidence intervals or variation across random seeds.

## 4 Discussion

### 4.1 Timing and Context

The consistent gain from adding RR timing, and the further gain from context for most families, indicate that explicit temporal and contextual inputs complement a short current-beat waveform in these pipelines. This agrees with the rationale of morphology-and-interval classifiers [6, 10] and of methods that supply RR information to learned representations [14–16]. A 255-sample window spans about 0.7 s and cannot contain the longer interval history that the RR features summarize. The gains cannot, however, be attributed solely to that information, because tuned settings, CNN capacity, and training duration also changed across stages.

The S4 descriptors add no new measurement: they are deterministic summaries of the waveform already supplied, so their value is representational. They helped Logistic Regression most, plausibly by exposing nonlinear waveform properties to a linear model, and slightly reduced Random Forest performance. Their class-level effects also differed by learner: Random Forest S, V, and F F1 decreased, XGBoost V and F F1 increased while S F1 decreased, and CNN F F1 increased while S F1 decreased. Summary features should therefore be evaluated per learner rather than assumed to help.

### 4.2 Sensitivity to Evaluation Choices

Three aspects of the evaluation changed which conclusions the same models supported. First, the choice of metric determined the ranking. The pipeline with the highest accuracy, Random Forest at S2, detected only 1.4% of S beats and none of the F beats, and the pipeline with the highest balanced accuracy, the CNN at S3, was not the one with the highest macro F1, XGBoost at S4. Second, development and evaluation disagreed on the best input stage for three of the four model families.

Third, and most important, minority-class performance on this benchmark depends on very few recordings. About 90% of DS1 fusion beats come from record 208 and 93% of DS2 fusion beats from record 213, so the F-class results mainly measure how well a model trained on the fusion beats of one subject recognizes those of another subject whose fusion morphology differs [22]. The record-level results confirm this. Record 213 supplied 203 of the 207 correct fusion detections made by the CNN, and outside that recording neither final-stage model detected more than 4 of the remaining 26 fusion beats. The precision–recall analysis further shows that the opposite fusion operating points of the two models lie on similar curves. The S-class results are likewise dominated by a single recording, since record 232 was almost entirely missed and excluding it nearly doubles S-class F1.

The bootstrap intervals show the consequence. When whole recordings are resampled, the 95% intervals for macro F1 are between 0.09 and 0.18 wide, and the gain from adding RR timing is the only change that is distinguishable from zero for every model family. These findings agree with earlier concerns that the composition of the MIT-BIH database constrains inter-patient training and evaluation [18]. They also imply that beat counts greatly overstate the effective sample size for the minority classes, because the evidence for each rare class comes from only one or two patients.

### 4.3 Comparison with Other Inter-Patient Results

Table 9 places the final-stage results alongside selected inter-patient results on the same DS1 and DS2 division. Published values are reproduced as reported; class sets, lead configurations, preprocessing, and operating points differ, so the comparison is indicative rather than matched. V-class F1 for final-stage XGBoost and the CNN, 0.926 to 0.954, is comparable to published values, but S-class F1, 0.199 to 0.227 at S4 and at most 0.245 at any stage, is far below the 0.51 to 0.73 reported elsewhere.

**Table 9:**
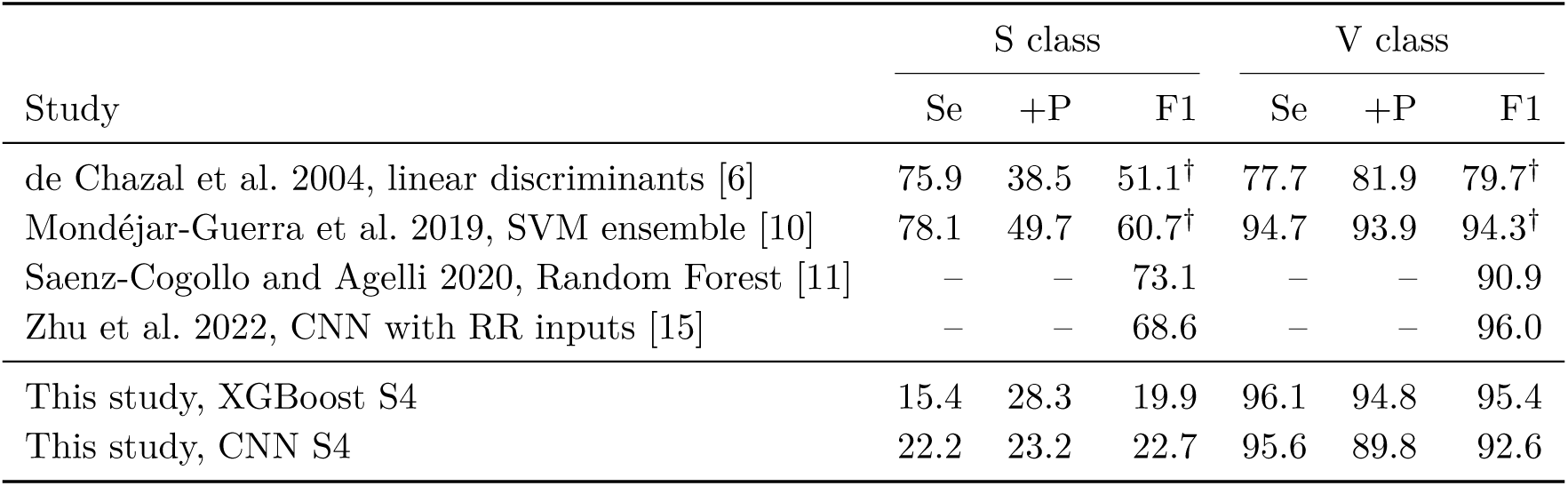
Selected inter-patient results on MIT-BIH DS2, in percent. Se denotes sensitivity, equivalent to recall, and +P denotes positive predictivity, equivalent to precision. Published values are taken directly from the original studies, and F1 values marked ^ψ^ were computed here from the reported Se and +P. Dashes mark values are not reported in the original study.

| Study | S class |  |  | V class |  |  |
| --- | --- | --- | --- | --- | --- | --- |
|  | Se | +P | F1 | Se | +P | F1 |
| de Chazal et al. 2004, linear discriminants [6] | 75.9 | 38.5 | 51.1<sup>†</sup> | 77.7 | 81.9 | 79.7<sup>†</sup> |
| Mondéjar-Guerra et al. 2019, SVM ensemble [10] | 78.1 | 49.7 | 60.7<sup>†</sup> | 94.7 | 93.9 | 94.3<sup>†</sup> |
| Saenz-Cogollo and Agelli 2020, Random Forest [11] | – | – | 73.1 | – | – | 90.9 |
| Zhu et al. 2022, CNN with RR inputs [15] | – | – | 68.6 | – | – | 96.0 |
| This study, XGBoost S4 | 15.4 | 28.3 | 19.9 | 96.1 | 94.8 | 95.4 |
| This study, CNN S4 | 22.2 | 23.2 | 22.7 | 95.6 | 89.8 | 92.6 |

The record-level results locate most of the S-class gap in one recording, and the feature definitions suggest why. Supraventricular beats are identified mainly by prematurity relative to the subject’s underlying rhythm and by atrial activity, while their QRS morphology is often normal. The present RR normalization divides each interval by the median of up to ten preceding intervals. In record 232, supraventricular beats outnumber all other beats by more than three to one, so that median is set largely by the supraventricular intervals themselves, and a premature beat no longer appears premature relative to it. This explanation was not tested directly, but it is consistent with the near-complete miss of record 232 alongside detection rates of 61 to 70% in the other recordings. RR intervals normalized by longer-term, subject-level averages have been used to reduce inter-subject differences in heart rate [9, 10] and would be a direct test. Per beat standardization also removes absolute amplitude, no P-wave specific features were extracted, and only one lead was used, whereas the reference study used two [6]. Finally, S prevalence doubled from 1.85% in DS1 to 3.70% in DS2, and the doubling comes almost entirely from record 232.

### 4.4 Limitations of the Study

The record-level bootstrap captures variability across evaluation patients but not across training runs, because each condition was trained only once with a single seed. Variation between seeds could widen the intervals further, particularly for the CNN, whose minority class predictions changed markedly between stages. Claims about training stability therefore require repeated training with several seeds per condition. Moreover, with only 22 evaluation recordings and the minority classes concentrated in two of them, the bootstrap intervals are themselves approximate.

The proposed mechanism for the failure on record 232 follows from the feature definitions but was not tested directly. Recomputing the RR features with subject-level normalization, or excluding ectopic intervals from the local median, would provide a direct test.

Because hyperparameters, training duration, and CNN capacity varied across stages, the stage effects are confounded with tuning. In addition, the S3 group combines rhythm-history and waveform-context features, and no baseline using the engineered features alone was evaluated. These contributions could be isolated by rerunning all stages with a fixed configuration, evaluating the 21 engineered features without the waveform, separating the two S3 feature groups, and including an unweighted control.

Several selected hyperparameters lay on the boundaries of their search grids, and search budgets differed across model families, which limits any ranking of the families. All reported class metrics use the default most-probable-class rule. The precision–recall analysis shows how strongly these metrics depend on the operating point, but no decision thresholds were tuned, since tuning them on DS2 would use evaluation labels for model selection.

The evaluation used one database with a known shared subject, records 201 and 202 [21]. Reference beat annotations, forward–backward filtering, post-fiducial samples, and the following RR interval make the task offline and annotation-assisted; beat detection errors, causal filtering, latency, and resource use were not evaluated, so the results do not establish real-time or wearable performance. Because DS2 results informed this analysis, any design changes they motivate require evaluation on independent data. Exact software versions were not archived with the reported outputs, which limits numerical reproducibility across environments.

## 5 Conclusion

Across three classical classifiers and a CNN, adding RR timing to a short beat waveform increased DS2 macro F1 for every model family, by 0.025 to 0.093. For all four models, the 95% record-level bootstrap interval for this gain lay entirely above zero, so the improvement was not an artifact of which recordings formed the test set. Broader context helped some families, while additional waveform summaries had model-dependent effects. These results support depend on evaluation choices. Accuracy, balanced accuracy, macro F1, and MCC ranked the pipelines differently. Development and evaluation favored the same stage for only one family. The final stage XGBoost and CNN could not be ranked, because their opposite operating points lie on similar precision–recall curves. Minority class performance rested on single recordings. Nearly all correct fusion detections came from record 213, and the weak supraventricular results reflect the near complete miss of record 232, plausibly because local RR normalization fails when ectopic beats dominate a recording. Studies using the MIT-BIH inter-patient division should therefore report record-level detections, record-level uncertainty, and threshold-independent metrics alongside aggregate scores.

## Data Availability

All data produced in the present study are available upon reasonable request to the authors.

https://physionet.org/content/mitdb/1.0.0/

## Acknowledgments

The authors thank the contributors and maintainers of the MIT-BIH Arrhythmia Database and PhysioNet for making the recordings available for research.

## Data and Code Availability

The MIT-BIH Arrhythmia Database, version 1.0.0, is publicly available through PhysioNet at https://physionet.org/content/mitdb/1.0.0/ [2]. The analysis was implemented in Python notebooks. Record partitions, fold assignments, selected settings, and feature definitions are given in the appendices. The DS2 predictions and class probabilities of all 16 final models, from which the record-level, bootstrap, and precision–recall analyses were computed, were exported from the saved models.

## A Record Partitions and Selected Settings

DS1: 101, 106, 108, 109, 112, 114, 115, 116, 118, 119, 122, 124, 201, 203, 205, 207, 208, 209, 215, 220, 223, 230.

DS2: 100, 103, 105, 111, 113, 117, 121, 123, 200, 202, 210, 212, 213, 214, 219, 221, 222, 228, 231, 232, 233, 234.

Validation folds: fold 1, records 114, 115, 119, 124, 203; fold 2, records 101, 109, 118, 201, 207; fold 3, records 106, 215, 223, 230; fold 4, records 108, 205, 209, 220; fold 5, records 112, 116, 122, 208.

**Table A.1:**
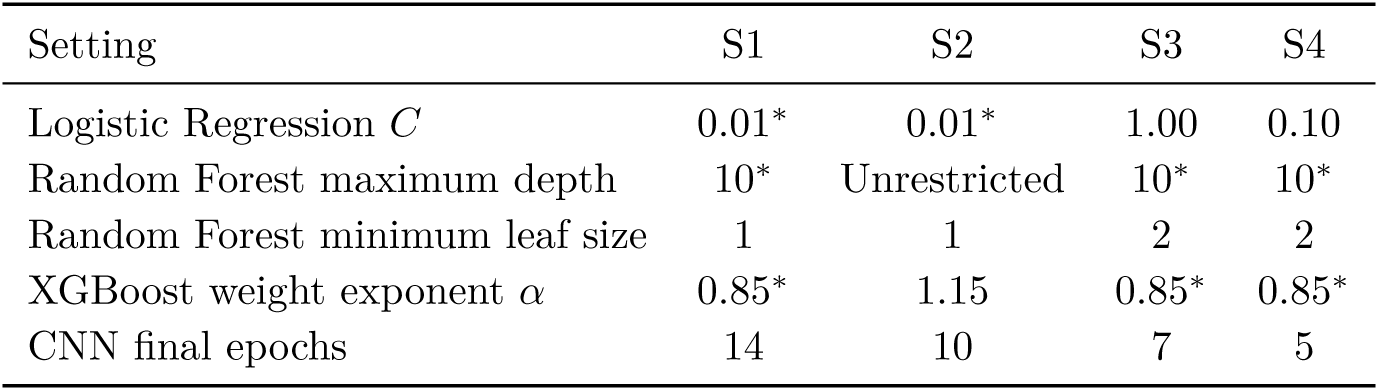
Selected settings by stage. Values marked ^∗^ indicate the lower boundary of the searched grid. The minimum leaf grid included only the values 1 and 2.

| Setting | S1 | S2 | S3 | S4 |
| --- | --- | --- | --- | --- |
| Logistic Regression $C$ | 0.01* | 0.01* | 1.00 | 0.10 |
| Random Forest maximum depth | 10* | Unrestricted | 10* | 10* |
| Random Forest minimum leaf size | 1 | 1 | 2 | 2 |
| XGBoost weight exponent $\alpha$ | 0.85* | 1.15 | 0.85* | 0.85* |
| CNN final epochs | 14 | 10 | 7 | 5 |

## B Development Results and Fold Composition

**Table B.1:** DS1 development macro F1, given as the mean *±* standard deviation across the five validation folds.

| Model | S1 | S2 | S3 | S4 |
| --- | --- | --- | --- | --- |
| Logistic Regression | 0.2769 ± 0.0887 | 0.3170 ± 0.0872 | 0.3806 ± 0.0608 | 0.4022 ± 0.0814 |
| Random Forest | 0.3796 ± 0.0618 | 0.4181 ± 0.0856 | 0.4338 ± 0.0502 | 0.4436 ± 0.0556 |
| XGBoost | 0.3779 ± 0.0957 | 0.4307 ± 0.0642 | 0.4679 ± 0.0465 | 0.4535 ± 0.0640 |
| CNN | 0.4174 ± 0.0814 | 0.5394 ± 0.1008 | 0.5528 ± 0.0798 | 0.5317 ± 0.0617 |

**Table B.2:** Size and minority class composition of the fixed DS1 validation folds.

| Fold | Records | Total beats | S beats | F beats |
| --- | --- | --- | --- | --- |
| 1 | 5 | 10,411 | 45 | 10 |
| 2 | 5 | 10,491 | 334 | 4 |
| 3 | 4 | 10,248 | 76 | 15 |
| 4 | 4 | 9,469 | 484 | 13 |
| 5 | 4 | 10,373 | 5 | 372 |

A note on the selection criterion: per-fold F1 on folds with four or five minority beats is highly unstable. Computing the selection criterion on pooled out-of-fold predictions would reduce this variance, although it would not remove the dependence between selection and the reported development score.

## C Per Class DS2 Results

In Tables C.1 to C.4, precision, recall, and F1 are proportions. Correct is the number of true beats of the class assigned to it, Predicted is the number of beats assigned to the class, and Support is the number of true beats of the class. Correct and predicted counts were computed from the exported DS2 predictions.

**Table C.1:** Logistic Regression per class DS2 results by stage.

| Stage | Class | Precision | Recall | F1 | Correct | Predicted | Support |
| --- | --- | --- | --- | --- | --- | --- | --- |
| S1 | N | 0.9322 | 0.6194 | 0.7442 | 27,401 | 29,395 | 44,241 |
|  | S | 0.0307 | 0.0887 | 0.0457 | 163 | 5,301 | 1,837 |
|  | V | 0.3418 | 0.5922 | 0.4335 | 1,907 | 5,579 | 3,220 |
|  | F | 0.0196 | 0.4742 | 0.0376 | 184 | 9,411 | 388 |
| S2 | N | 0.9632 | 0.7201 | 0.8241 | 31,859 | 33,077 | 44,241 |
|  | S | 0.1263 | 0.1753 | 0.1468 | 322 | 2,550 | 1,837 |
|  | V | 0.5072 | 0.7488 | 0.6047 | 2,411 | 4,754 | 3,220 |
|  | F | 0.0261 | 0.6263 | 0.0501 | 243 | 9,305 | 388 |
| S3 | N | 0.9581 | 0.7847 | 0.8628 | 34,715 | 36,232 | 44,241 |
|  | S | 0.0860 | 0.1568 | 0.1110 | 288 | 3,350 | 1,837 |
|  | V | 0.6484 | 0.9025 | 0.7546 | 2,906 | 4,482 | 3,220 |
|  | F | 0.0390 | 0.5644 | 0.0729 | 219 | 5,622 | 388 |
| S4 | N | 0.9590 | 0.8629 | 0.9084 | 38,174 | 39,804 | 44,241 |
|  | S | 0.0982 | 0.1889 | 0.1292 | 347 | 3,533 | 1,837 |
|  | V | 0.7952 | 0.8997 | 0.8442 | 2,897 | 3,643 | 3,220 |
|  | F | 0.0484 | 0.3376 | 0.0847 | 131 | 2,706 | 388 |

**Table C.2:** Random Forest per class DS2 results by stage.

| Stage | Class | Precision | Recall | F1 | Correct | Predicted | Support |
| --- | --- | --- | --- | --- | --- | --- | --- |
| S1 | N | 0.9439 | 0.9510 | 0.9475 | 42,075 | 44,575 | 44,241 |
|  | S | 0.0461 | 0.0419 | 0.0439 | 77 | 1,669 | 1,837 |
|  | V | 0.8060 | 0.8152 | 0.8106 | 2,625 | 3,257 | 3,220 |
|  | F | 0.0000 | 0.0000 | 0.0000 | 0 | 185 | 388 |
| S2 | N | 0.9431 | 0.9988 | 0.9702 | 44,190 | 46,858 | 44,241 |
|  | S | 0.7879 | 0.0142 | 0.0278 | 26 | 33 | 1,837 |
|  | V | 0.9714 | 0.8429 | 0.9026 | 2,714 | 2,794 | 3,220 |
|  | F | 0.0000 | 0.0000 | 0.0000 | 0 | 1 | 388 |
| S3 | N | 0.9539 | 0.9862 | 0.9698 | 43,631 | 45,739 | 44,241 |
|  | S | 0.3178 | 0.0925 | 0.1433 | 170 | 535 | 1,837 |
|  | V | 0.9022 | 0.9450 | 0.9231 | 3,043 | 3,373 | 3,220 |
|  | F | 0.1538 | 0.0155 | 0.0281 | 6 | 39 | 388 |
| S4 | N | 0.9517 | 0.9881 | 0.9695 | 43,713 | 45,932 | 44,241 |
|  | S | 0.3214 | 0.0827 | 0.1316 | 152 | 473 | 1,837 |
|  | V | 0.9122 | 0.9227 | 0.9174 | 2,971 | 3,257 | 3,220 |
|  | F | 0.1250 | 0.0077 | 0.0146 | 3 | 24 | 388 |

**Table C.3:** XGBoost per class DS2 results by stage.

| Stage | Class | Precision | Recall | F1 | Correct | Predicted | Support |
| --- | --- | --- | --- | --- | --- | --- | --- |
| S1 | N | 0.9557 | 0.9190 | 0.9370 | 40,659 | 42,543 | 44,241 |
|  | S | 0.0855 | 0.0789 | 0.0821 | 145 | 1,696 | 1,837 |
|  | V | 0.5878 | 0.8963 | 0.7100 | 2,886 | 4,910 | 3,220 |
|  | F | 0.0093 | 0.0129 | 0.0108 | 5 | 537 | 388 |
| S2 | N | 0.9628 | 0.9094 | 0.9353 | 40,231 | 41,785 | 44,241 |
|  | S | 0.2330 | 0.1644 | 0.1928 | 302 | 1,296 | 1,837 |
|  | V | 0.8502 | 0.9602 | 0.9019 | 3,092 | 3,637 | 3,220 |
|  | F | 0.0179 | 0.1366 | 0.0316 | 53 | 2,968 | 388 |
| S3 | N | 0.9597 | 0.9773 | 0.9684 | 43,238 | 45,055 | 44,241 |
|  | S | 0.2914 | 0.1551 | 0.2025 | 285 | 978 | 1,837 |
|  | V | 0.9209 | 0.9578 | 0.9390 | 3,084 | 3,349 | 3,220 |
|  | F | 0.1908 | 0.1495 | 0.1676 | 58 | 304 | 388 |
| S4 | N | 0.9591 | 0.9791 | 0.9690 | 43,317 | 45,165 | 44,241 |
|  | S | 0.2831 | 0.1535 | 0.1991 | 282 | 996 | 1,837 |
|  | V | 0.9479 | 0.9609 | 0.9543 | 3,094 | 3,264 | 3,220 |
|  | F | 0.2452 | 0.1649 | 0.1972 | 64 | 261 | 388 |

**Table C.4:**
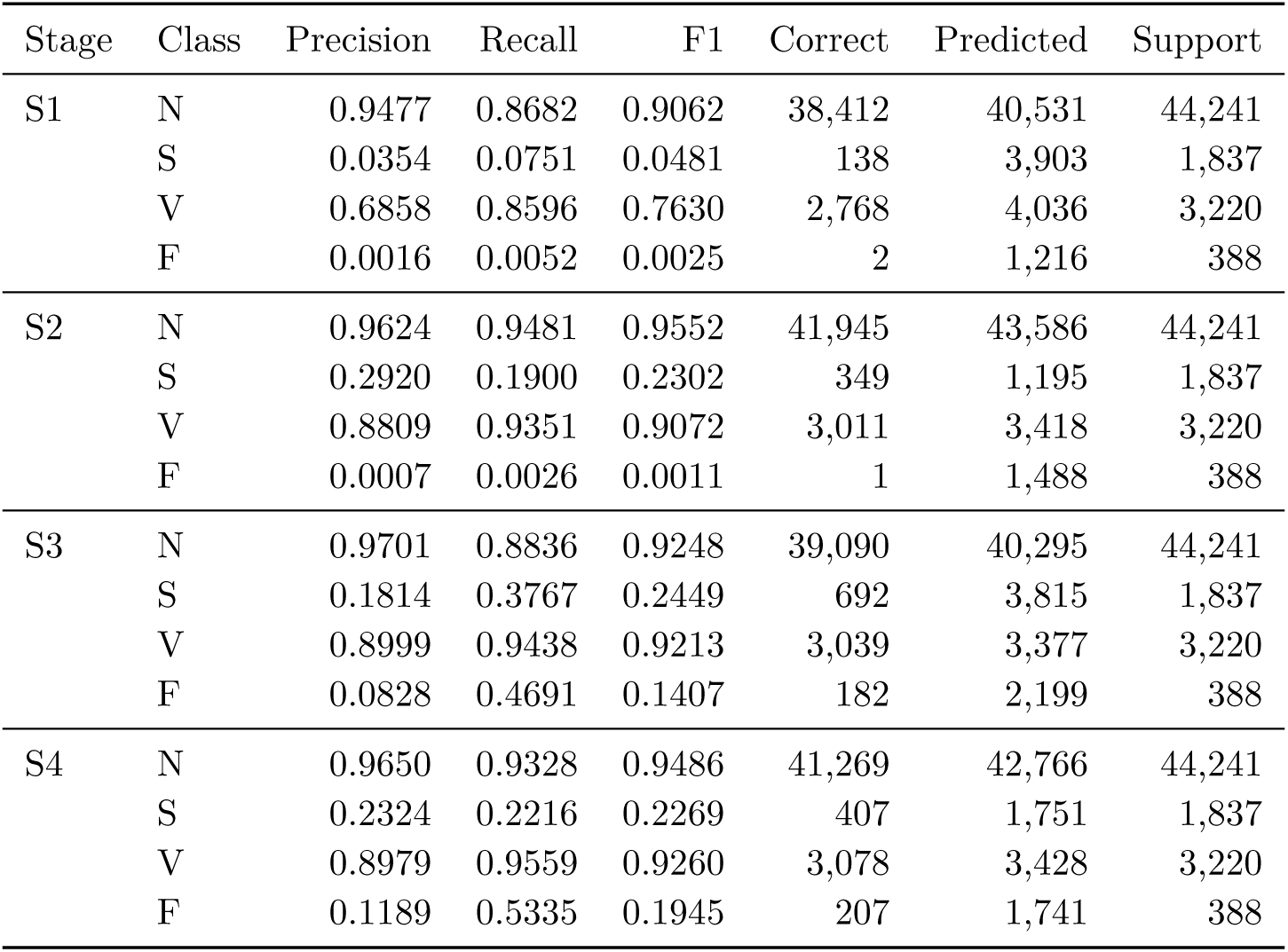
CNN per class DS2 results by stage.

| Stage | Class | Precision | Recall | F1 | Correct | Predicted | Support |
| --- | --- | --- | --- | --- | --- | --- | --- |
| S1 | N | 0.9477 | 0.8682 | 0.9062 | 38,412 | 40,531 | 44,241 |
|  | S | 0.0354 | 0.0751 | 0.0481 | 138 | 3,903 | 1,837 |
|  | V | 0.6858 | 0.8596 | 0.7630 | 2,768 | 4,036 | 3,220 |
|  | F | 0.0016 | 0.0052 | 0.0025 | 2 | 1,216 | 388 |
| S2 | N | 0.9624 | 0.9481 | 0.9552 | 41,945 | 43,586 | 44,241 |
|  | S | 0.2920 | 0.1900 | 0.2302 | 349 | 1,195 | 1,837 |
|  | V | 0.8809 | 0.9351 | 0.9072 | 3,011 | 3,418 | 3,220 |
|  | F | 0.0007 | 0.0026 | 0.0011 | 1 | 1,488 | 388 |
| S3 | N | 0.9701 | 0.8836 | 0.9248 | 39,090 | 40,295 | 44,241 |
|  | S | 0.1814 | 0.3767 | 0.2449 | 692 | 3,815 | 1,837 |
|  | V | 0.8999 | 0.9438 | 0.9213 | 3,039 | 3,377 | 3,220 |
|  | F | 0.0828 | 0.4691 | 0.1407 | 182 | 2,199 | 388 |
| S4 | N | 0.9650 | 0.9328 | 0.9486 | 41,269 | 42,766 | 44,241 |
|  | S | 0.2324 | 0.2216 | 0.2269 | 407 | 1,751 | 1,837 |
|  | V | 0.8979 | 0.9559 | 0.9260 | 3,078 | 3,428 | 3,220 |
|  | F | 0.1189 | 0.5335 | 0.1945 | 207 | 1,741 | 388 |

## D Threshold Independent Results and Confusion Matrices

**Table D.1:** Average precision on DS2 for each class, model, and stage, computed from the class probabilities by comparing each class against all other classes.

| Model | Stage | N | S | V | F |
| --- | --- | --- | --- | --- | --- |
| Logistic Regression | S1 | 0.918 | 0.031 | 0.427 | 0.016 |
|  | S2 | 0.972 | 0.090 | 0.615 | 0.022 |
|  | S3 | 0.947 | 0.075 | 0.768 | 0.029 |
|  | S4 | 0.941 | 0.085 | 0.850 | 0.027 |
| Random Forest | S1 | 0.966 | 0.043 | 0.861 | 0.014 |
|  | S2 | 0.986 | 0.126 | 0.957 | 0.018 |
|  | S3 | 0.976 | 0.111 | 0.973 | 0.106 |
|  | S4 | 0.968 | 0.111 | 0.972 | 0.040 |
| XGBoost | S1 | 0.983 | 0.048 | 0.895 | 0.012 |
|  | S2 | 0.982 | 0.134 | 0.976 | 0.025 |
|  | S3 | 0.985 | 0.147 | 0.985 | 0.121 |
|  | S4 | 0.977 | 0.147 | 0.991 | 0.139 |
| CNN | S1 | 0.962 | 0.038 | 0.869 | 0.014 |
|  | S2 | 0.954 | 0.128 | 0.962 | 0.017 |
|  | S3 | 0.981 | 0.173 | 0.980 | 0.067 |
|  | S4 | 0.986 | 0.203 | 0.985 | 0.151 |

**Table D.2:**
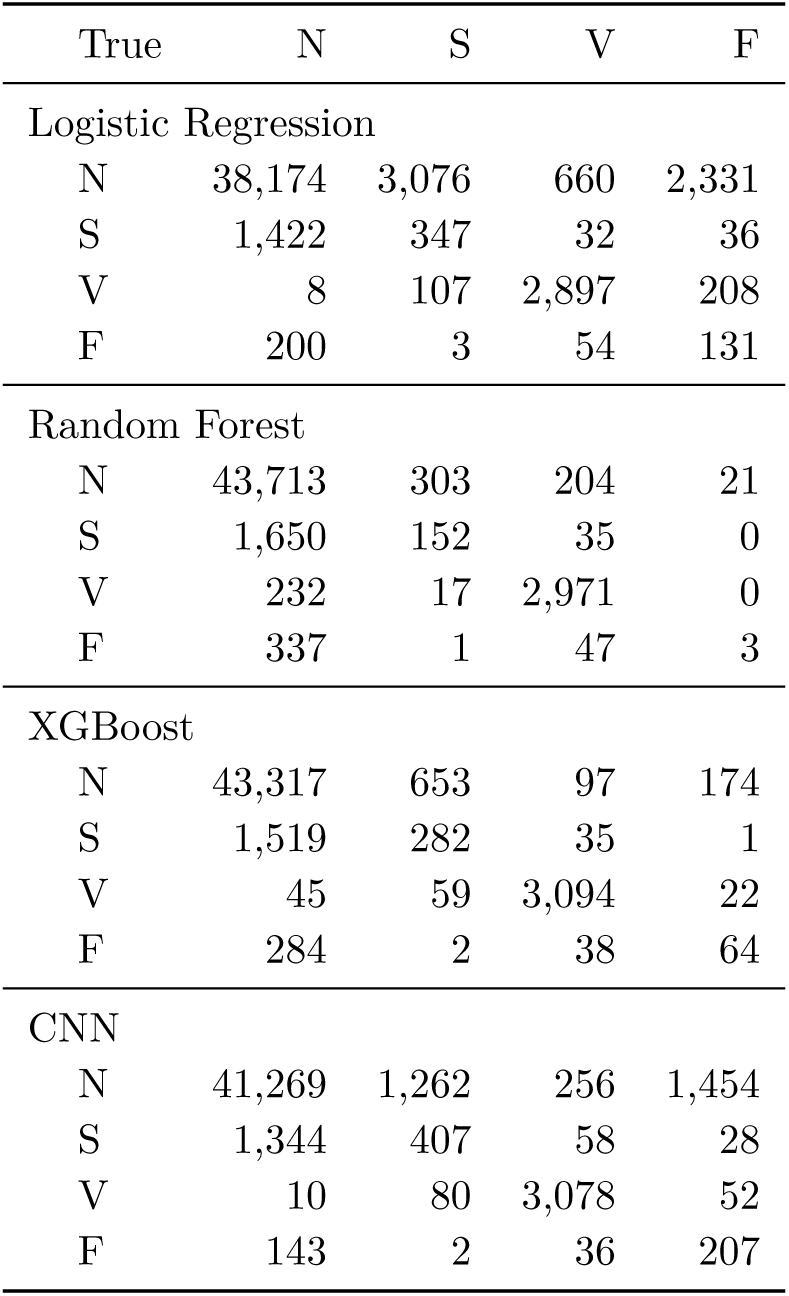
DS2 confusion matrices for the final-stage S4 models. Rows are true classes and columns are predicted classes.

## E Feature Definitions

Let *t_i_* be the time in seconds of the current beat event and *x_i_ ∈* R*^L^* its standardized waveform, with *L* = 255. The six RR features are

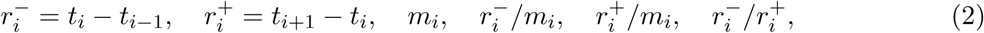

where *m_i_* is the median of up to ten intervals preceding 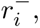 namely 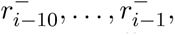 with at least three required. The historical mean *µ_i_*, population standard deviation *σ_i_*, and coefficient of variation *σ_i_/µ_i_*of the same intervals are the rhythm-history descriptors. Missing neighbors or insufficient history produce missing values, and ratios require finite inputs and positive denominators.

For waveform context, the references are the immediately preceding beat waveform, when available, and the pointwise median of available waveforms among the five preceding beat positions, which requires at least three waveforms; invalid positions are not replaced by more distant beats. For each reference *z*, the Pearson correlation and root-mean-square distance are

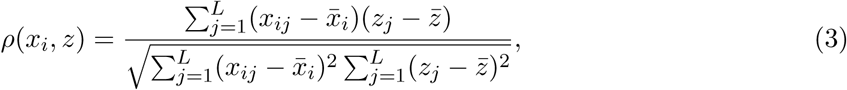

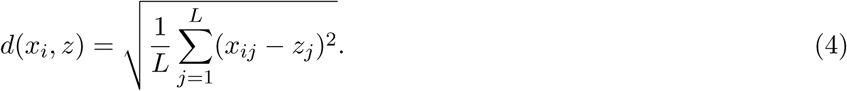

Unavailable or unequal-length references give missing values. Correlation additionally requires both waveform standard deviations to be at least 10*^−^*^8^. The median template is not restandardized after construction.

For the eight current-waveform descriptors, let *x̄* = *L^−^*^1^Σ*_j_ x_j_* and *m*_k_ = *L^−^*^1^ (*x − x̄*)*^k^*. The amplitude descriptors are min*_j_ x_j_*, max*_j_ x_j_*, their difference, and *L^−^*^1^Σ*_j_ |x_j_|*. Bias-corrected skewness and excess kurtosis, computed with SciPy using bias=False, with fisher=True for kurtosis [29], are

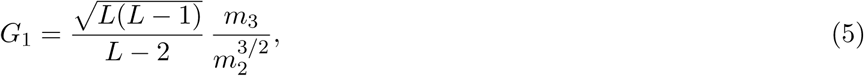

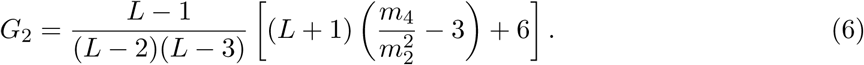

The remaining descriptors are 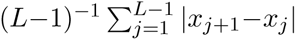 and max*_j_ |x_j_*_+1_*−x_j_|*. They are dimensionless sample differences, not voltage derivatives per second.

The features are not statistically independent. For two nonconstant waveforms each standardized to zero mean and unit population variance, *d*^2^ = 2(1 *− ρ*); this identity applies to the preceding-beat comparison up to numerical precision but need not apply to the unstandardized median template. Peak-to-peak amplitude is likewise determined by the minimum and maximum. The feature count therefore overstates the number of independent information sources.

As an illustration of overfitting within a fold, for the S4 CNN in validation fold 2 the training loss decreased from 0.4646 at epoch 1 to 0.0764 at epoch 20 while validation macro F1 decreased from 0.4764 to 0.3685. This single trajectory motivates checkpoint selection but does not summarize all folds.

